# Evaluation of the clinical characteristics of suspected or confirmed cases of COVID-19 during home care with isolation: A new retrospective analysis based on O2O

**DOI:** 10.1101/2020.02.26.20028084

**Authors:** Hui Xu, Sufang Huang, Shangkun Liu, Juan Deng, Bo Jiao, Ling Ai, Yaru Xiao, Li Yan, Shusheng Li

## Abstract

**Background:** The recent outbreak of the novel coronavirus in December 2019 (COVID-19) has activated top-level response nationwide. We developed a new treatment model based on the online-to-offline (O2O) model for the home isolated patients, because in the early stages the medical staff were insufficient to cope with so many patients.

**Methods:** In this single-centered, retrospective study, we enrolled 48 confirmed/suspected COVID-19 patients who underwent home isolation in Wuhan between January 6 and January 31, 2020. By WeChat and online document editing all patients were treated with medical observation scale. The clinical indications such as Fever, Muscle soreness, Dyspnea and Lack of strength were collected with this system led by medical staff in management, medicine, nursing, rehabilitation and psychology.

**Findings:** The mean age of 48 patients was 39·08±13·88 years, 35(72·9%) were women. Compared with non-hospitalized patients, inpatients were older(≥70years, 2·4% vs 33·3%, P<0·04). All inpatients had fever, 50% inpatients had coughs and showed infiltration in both lungs at the time of diagnosis. 33·3% inpatients exhibited negative changes in their CT results at initial diagnosis. The body temperature of non-hospitalized patients with mild symptoms returned to normal by day 4-5. While dyspnea peaked on day 6 for non-hospitalized patients with mild symptoms, it persisted in hospitalized patients and exacerbated over time. The lack of strength and muscle soreness were both back to normal by day 4 for non-hospitalized patients.

**Interpretation:** Monitoring the trends of symptoms is more important for identifying severe cases. Excessive laboratory data and physical examination are not necessary for the evaluation of patients with mild symptoms. The system we developed is the first to convert the subjective symptoms of patients into objective scores. This type of O2O, subjective-to-objective strategy may be used in regions with similar highly infectious diseases to minimize the possibility of infection among medical staff.

## Introduction

The recent outbreak of the novel coronavirus in December 2019 (COVID-19) has activated top-level response nationwide and has been classified as a public health emergency of international concern (PHIEC) by the World Health Organization (WHO).^1^ By 24:00 on February 16, 2020, 70,548 confirmed cases, 10,644 severe cases, 1,770 deaths, and 546,016 close contact cases have been identified in 31 provinces (autonomous regions and municipalities) and the Xinjiang Production and Construction Corps of China.^2^ The SARS-CoV-2-induced pneumonia has rapidly spread from Wuhan to 21 other countries, including the United States, Japan, Italy and Germany^3,4^, demonstrating high levels of infectivity and pathogenicity.^5-7^

The fever clinic of Tongji hospital in Wuhan has been the center of this outbreak. During the early phase of this outbreak, a large number of patients poured into the fever clinic, which far exceeded the medical resources that the hospital could equip. The medical staff were obviously insufficient to cope with it, and were prone to a wide range of cross-infection between doctors and patients. Based on this situation many patients had to be quarantined at home due to objective reasons and could not receive effective medical guidance. Therefore, we developed a new treatment method based on the online-to-offline (O2O) business model^8^ for close contact, suspected (currently known as clinical diagnosis) and confirmed patients that were under quarantine. We developed a medical observation scale according to the patients’ first symptoms and new symptoms which can be filled out by the patients on their smartphones or computers. Our online multidisciplinary team (medicine, rehabilitation, psychology and nursing) can then provide guidance and advice for patients based on the subjective changes in their symptoms. This method ensures that the patients follow an orderly treatment-seeking strategy that begins from their home and extends to the community and finally to the hospital. Our strategy not only helps relief the problem of scarce medical resources and reduce unnecessary cross-infection in hospitals, but it also increases people’s self-management ability and cooperation and encourages them to participate in health monitoring. In addition, in accordance with the guidelines of home care for isolation patients issued by WHO^9^, the infection of family members was not increased during the strict self-isolation at home in this study.

In this study, we reviewed the observational data of isolated patients and examined the temporal relationship among their clinical symptoms in order to help physicians in areas with insufficient medical resources to effectively identify and treat critical patients.

## Research in context

### Evidence before this study

Since December 2019, COVID-19 has exploded in Wuhan, China. So far, it has spread all over the world. At the initial stage of the outbreak of this new type of respiratory infectious disease, due to the obvious shortage of medical resources, a large number of patients had to be isolated and treated at home. Up to Feb 24, 2020, we have searched PubMed for all the literatures related to COVID-19. There is no description of the clinical characteristics and outcome of patients during home isolation. In addition, we have created a medical observation scale designed for patients quarantined at home. Combined with the O2O model the patients were evaluated and treated.

### Added value of this study

We report the clinical characteristics and outcome of 48 confirmed/suspected COVID-19 patients from 188 individuals who participated in the medical observation during home care and isolation. Compared with non-hospitalized patients, inpatients were older(≥70years, 2·4% vs 33·3%, P<0·04). All inpatients had fever, 50% inpatients had coughs and showed infiltration in both lungs by at the time of diagnosis. 33·3% inpatients exhibited negative changes in their CT results at initial diagnosis. The body temperature of non-hospitalized patients with mild symptoms returned to normal by day 4-5. While dyspnea peaked on day 6 for non-hospitalized patients with mild symptoms, it persisted in hospitalized patients and exacerbated over time. The lack of strength and muscle soreness were both back to normal by day 4 for non-hospitalized patients.

### Implications of all the available evidence

Monitoring the trends of symptoms is more important for identifying severe cases. Excessive laboratory data and physical examination are not necessary for the evaluation of patients with mild symptoms. The system we developed is the first to convert the subjective symptoms of patients into objective scores. This type of O2O, subjective-to-objective strategy may prove useful in areas with insufficient medical resources to minimize the possibility of infection among patients and medical staff.

## Methods

### Study design and inclusion criteria

This is a single-center retrospective study of individuals who came into close contacts with COVID-19 patients in Tongji hospital of Wuhan between January 6 and January 31, 2020. The study was approved by Tongji Hospital Ethics Committee before data were collected retrospectively. Among the 188 individuals who participated in the medical observation, 114 were excluded from this study due to no signs of discomfort during the 14-day quarantine (108 cases) and non-COVID-19 (6 cases). Among the 74 confirmed/suspected COVID-19 cases, 26 were removed due to incomplete data, and a final total of 48 confirmed/suspected cases were included in this retrospective study.

### Inclusion criteria

Patients who were confirmed with or suspected of COVID-19 and were willing to undergo medical observation.

Diagnostic criteria^10^:

1. Epidemiological history: Traveled or lived in Wuhan within 14 days before onset; Had contact with patients with fever and respiratory symptoms from Wuhan within 14 days before onset; Had contact with COVID-19 patients (positive for COVID-19 nucleic acid) within 14 days before onset; Or part of a familial cluster of onset;
2. Clinical manifestations: Fever and/or respiratory symptoms; Normal or decreased total white blood cell count or decreased lymphocyte count during early stage of onset; Typical imaging features;

Subjects that meet any one epidemiological history or meet two clinical manifestations without epidemiological history were defined as suspected cases.

Confirmed case is defined as a suspected case with one of the following etiological evidence: Positive for SARS-CoV-2 nucleic acid in respiratory or blood samples detected by RT-PCR; Virus sequence detected in respiratory or blood samples shares high homology with the known sequence of SARS-CoV-2.

### Exclusion criteria

1. Pregnant or breast-feeding women;
2. <18 or >75 years old;
3. Unable to cooperate with data reporting.

### Endpoints

1. The end date of the observation was January 31, 2020;
2. When the patient was admitted to the hospital or died;
3. When the patient was clinically cured: with normal CT imaging and at least twice of SARS-CoV-2 nucleic acid assays were negative.

### Medical observation scales

1. Fever: 1 = None (37·3◻ and below); 2 = Low grade fever (37·3∼38◻); 3 = Moderate fever (38·1∼39◻); 4 = High fever (39·1∼40◻); 5 = Hyperpyrexia (40◻ and higher)
2. Mental state: 1 = Good; 2 = Average; 3 = Poor
3. Muscle soreness: 1 = Complete absence of soreness; 2 = Light pain felt only when touched; 3 = Occasional soreness; 4 = Sustained soreness
4. Cough: 1 = None; 2 = Occasional; 3 = Frequent and slightly interferes with daily activities; 4 = Frequent and seriously interferes with daily activities
5. Dyspnea: 1 = Not troubled by breathlessness except with strenuous exercise; 2 = Troubled by shortness of breath when hurrying on a level surface or walking up a slight hill; 3 = Walks slower than normal based on age on a level surface due to breathlessness or has to stop for breath when walking on level surface at own pace; 4 = Stops for breath after walking 100 meters or after a few minutes on a level surface; 5 = Too breathless to leave the house
6. Lack of strength: 1 = No lack of strength; 2 = Mild: Slightly lacked strength, able to do physical work, improves after rest but does not recover to normal; 3 = Moderate: Lacked strength, feels weak, able to persist in daily activities and work but light physical work is very tiring and does not recover to normal after long periods of rest; 4 = Severe: Extremely lacked strength, unable to conduct normal activities, feels tired at rest, cannot talk
7. Diarrhea: 1 = No diarrhea; 2 = Mild diarrhea: Loose stool for no more than 3 times; 3 = Moderate diarrhea: 4-6 times; 4 = Severe diarrhea: More than 6 times
8. Chest tightness: 1 = None; 2 = Mild; 3 = Moderate; 4 = Severe

### Development of medical observation scale

We first obtained the corresponding symptoms from the 6 subjects under observation, then continuously added new symptoms to the online scale as we furthered our understanding of the disease. We graded the symptoms based on our clinical work experience in lay language and compiled them into the medical observation scale. We gave our scale to 34 experts for evaluation of the items and score criteria.

Two rounds of expert consultations were conducted with 17 experts from the emergency department, respiratory department, intensive care unit, and infectious disease department each round. Statistical analysis of the consultation results showed that the response rates of the two rounds of expert consultation were 100% and 88·24%, and the mean authoritative coefficient was 0·855. The degree of coordination of expert opinions (W value) was 0·204 and 0·293 for the two rounds, respectively, and the W values were statistically significant (P<0·01). The levels of enthusiasm and authority, concentration of opinion, and consistency in item evaluation results were relatively high among the experts, which demonstrates that the “Quarantine Management Assessment Form for Suspected or Confirmed COVID-19 Patients with Mild Symptoms” we designed was scientifically sound and reliable and can be used for the centralized quarantine, observation and care of suspected or confirmed cases. This assessment form includes general demographics, past history, current medical history, current symptoms and signs, and diagnostic recommendations consisting of 5 primary indicators, 22 secondary indicators, and 83 tertiary indicators.

### Treatment

Empirical therapy consisted of oral moxifloxacin or levofloxacin (consider tolerance) and arbidol. Arbidol was approved in China and Russia for influenza treatment. In vitro studies showed that arbidol had inhibitory effects on SARS.^11^

### Statistical analysis

Statistical analysis was performed using SPSS26·0. Normally distributed measured data were expressed as mean±standard deviation (x±s) and compared using the t-test based on homogeneity of variance. Categorical data were expressed as frequency and percentage and compared using the χ^2^ test. P<0·05 was considered statistically significant.

## Results

### 1. Demographics, baseline characteristics, and clinical outcomes of 48 COVID-19 patients quarantined at home (Table 1)

**Table 1:**
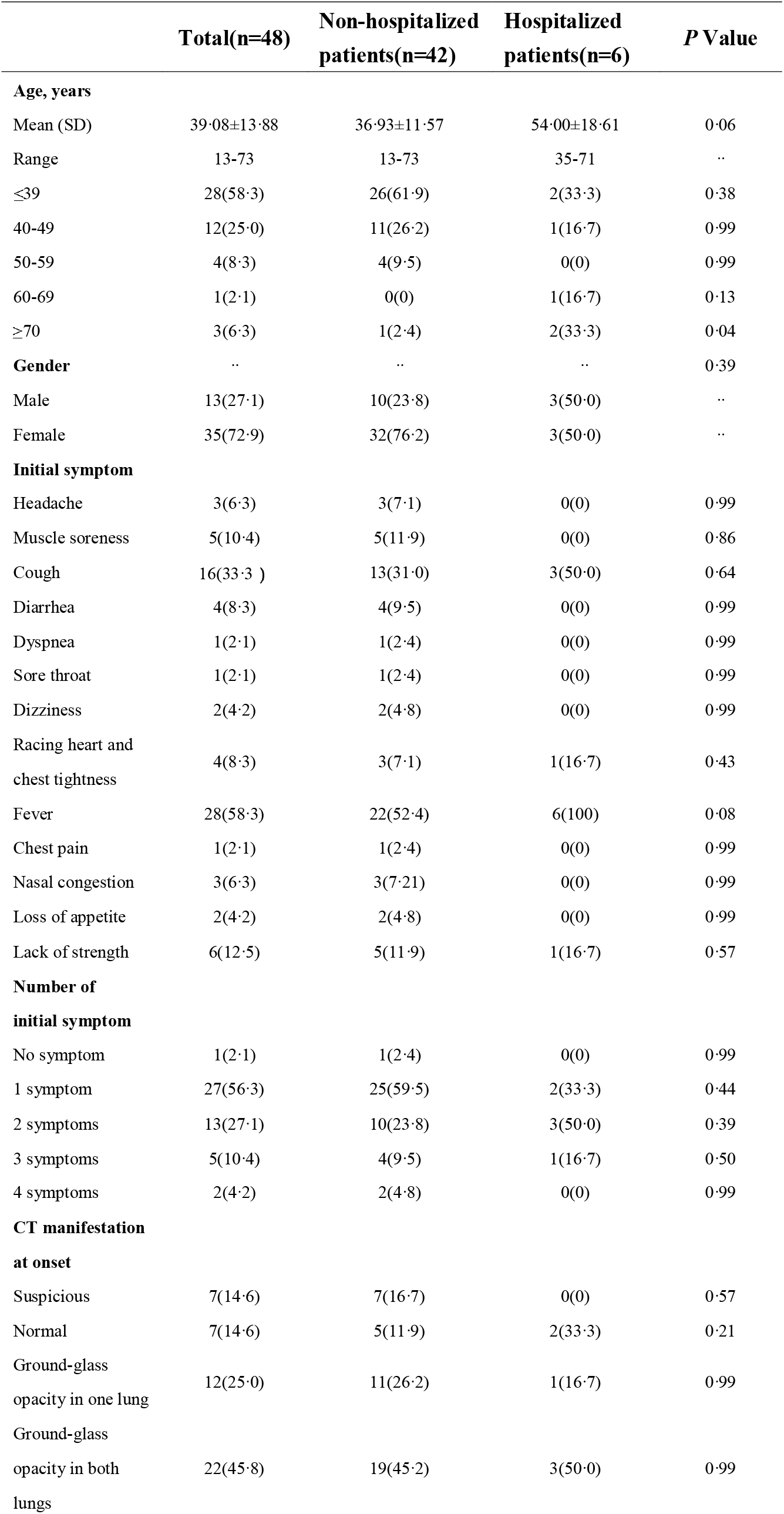
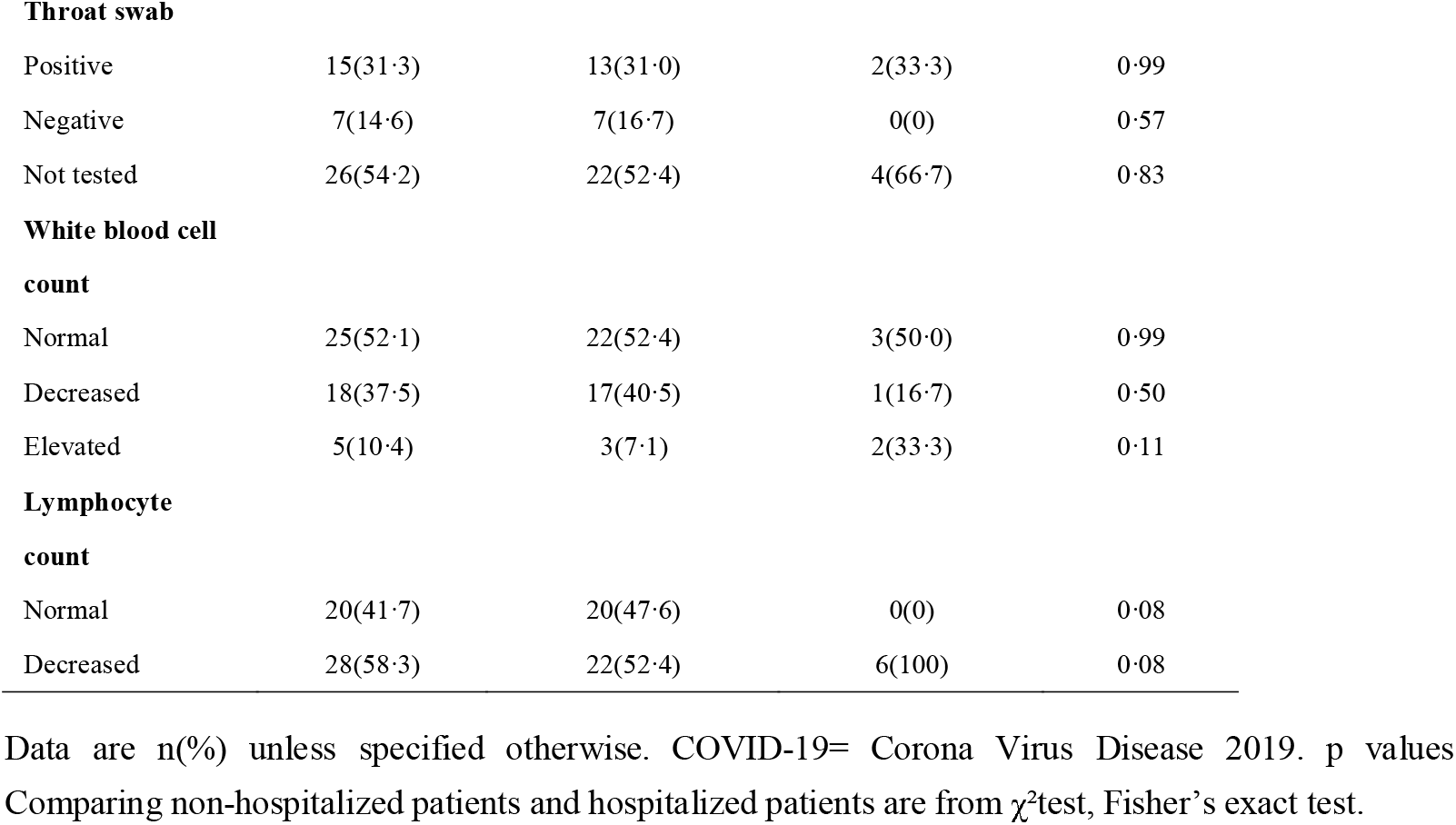
Demographics, baseline characteristics, and clinical outcomes of 48 COVID-19 patients quarantined at home.

The mean age of the 48 patients was 39·08±13·88 years. The patients were predominantly young and middle-aged adults, with 72·9% being females. Fever is the most common initial symptom (58·3%), followed by cough (33·3%), lack of strength (12·5%), and muscle soreness (10·4%). Contrary to previous reports, 3 patients (6·3%) had nasal congestion and 1 patient (2·1%) lacked symptoms during screening.

There were 13 patients with two initial symptoms, 5 patients with three initial symptoms, and 2 patients (4·2%) with four initial symptoms. CT results were normal for 14·6% of patients during initial diagnosis, which increased the difficulty of clinical screening.

The percentage of patients hospitalized during the observation was significantly higher among elderly patients (2·4% vs 33·3%, P<0·04). All inpatients (100%) had fever, 50% patients had coughs, and 50% patients showed infiltration in both lungs by at the time of diagnosis. One-third of the critical patients (33·3%) exhibited negative changes in their CT results at initial diagnosis.

### 2. Clinical characteristics of 6 patients admitted to hospital (Table 2)

**Table 2.**
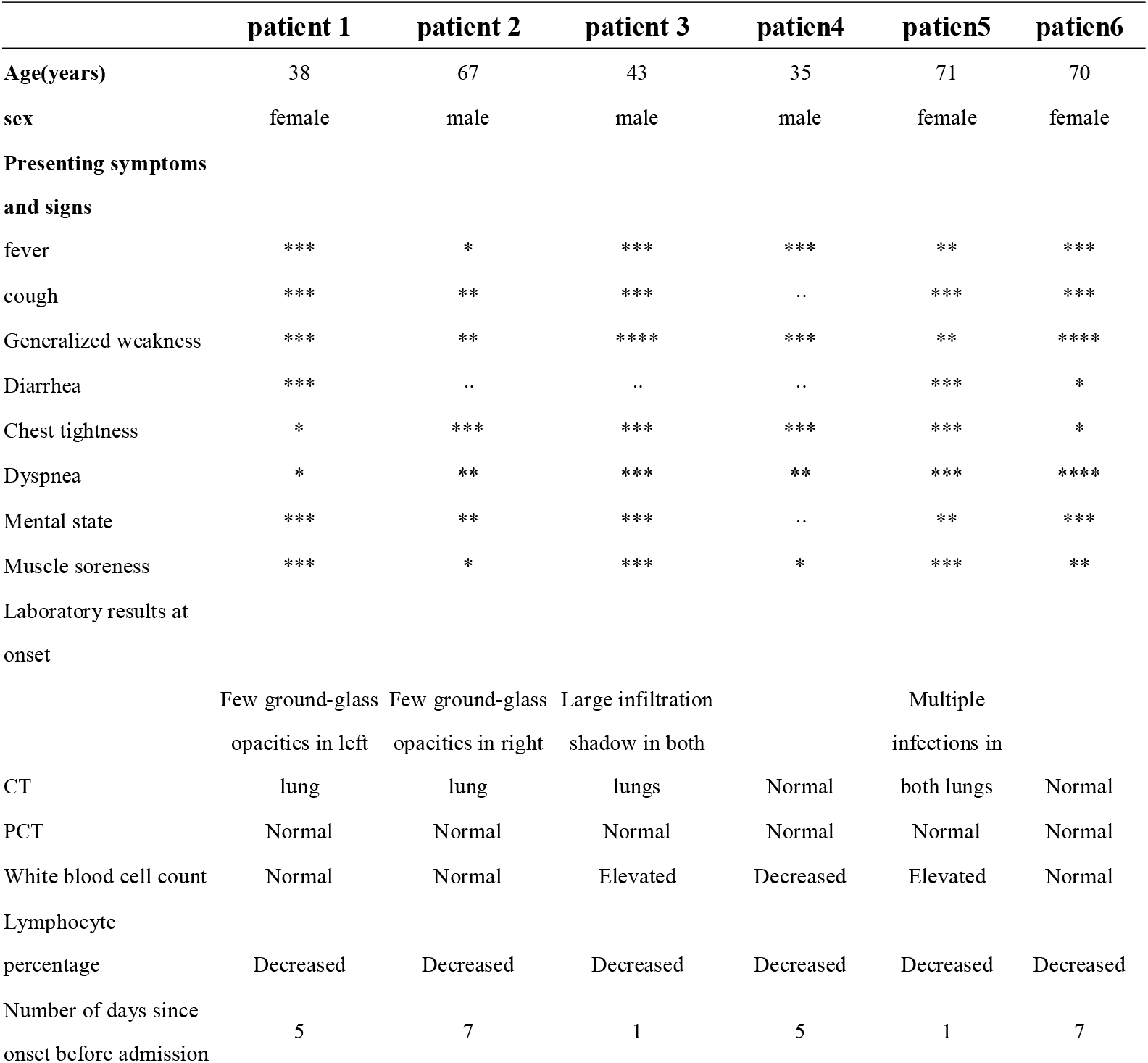
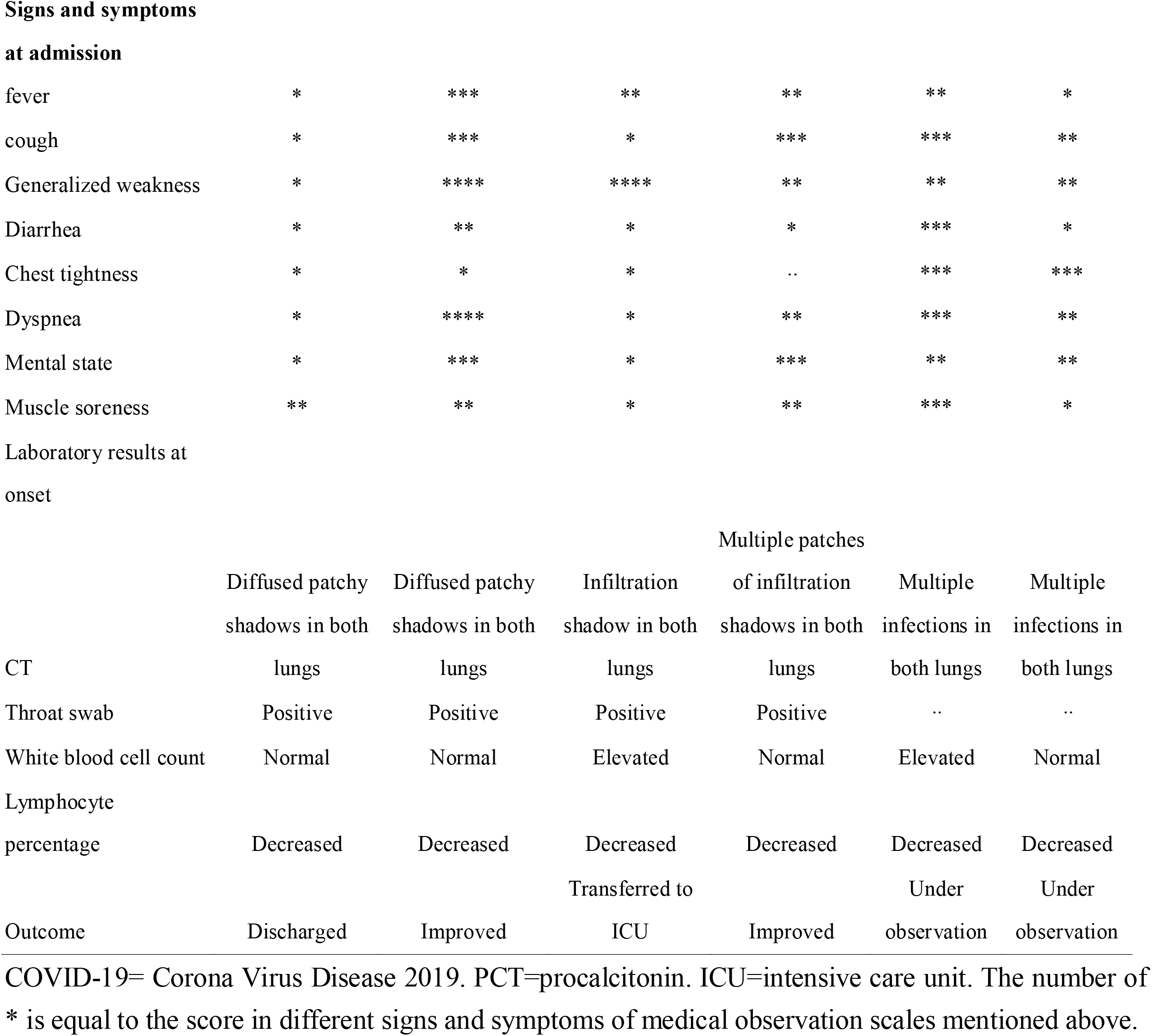
Clinical characteristics of 6 patients admitted to hospital.

Among the 6 patients who were admitted to our hospital, 1 patient (Patient 3) was found to be in critical conditions and was directly admitted to the ICU. This patient no longer required invasive ventilation at the time of manuscript submission.

Patient 5 was our first patient. She was found to have persistent fever on day 5 of observation and her CT results indicated progression. Although the clinical information of the patient was not disclosed at the time, we observed that the lack of strength, chest tightness and dyspnea the patient developed did not match her age and CT changes. Therefore, the patient was admitted into our hospital for treatment. The patient’s conditions deteriorated as disease progressed, and both her lungs appeared as “white lungs” (figure 2). Noninvasive ventilation was provided to the patient and her conditions gradually improved. The patient’s SPO_2_ is currently around 95% on room air and she is able to get off the hospital bed by herself.

**Figure 1.**
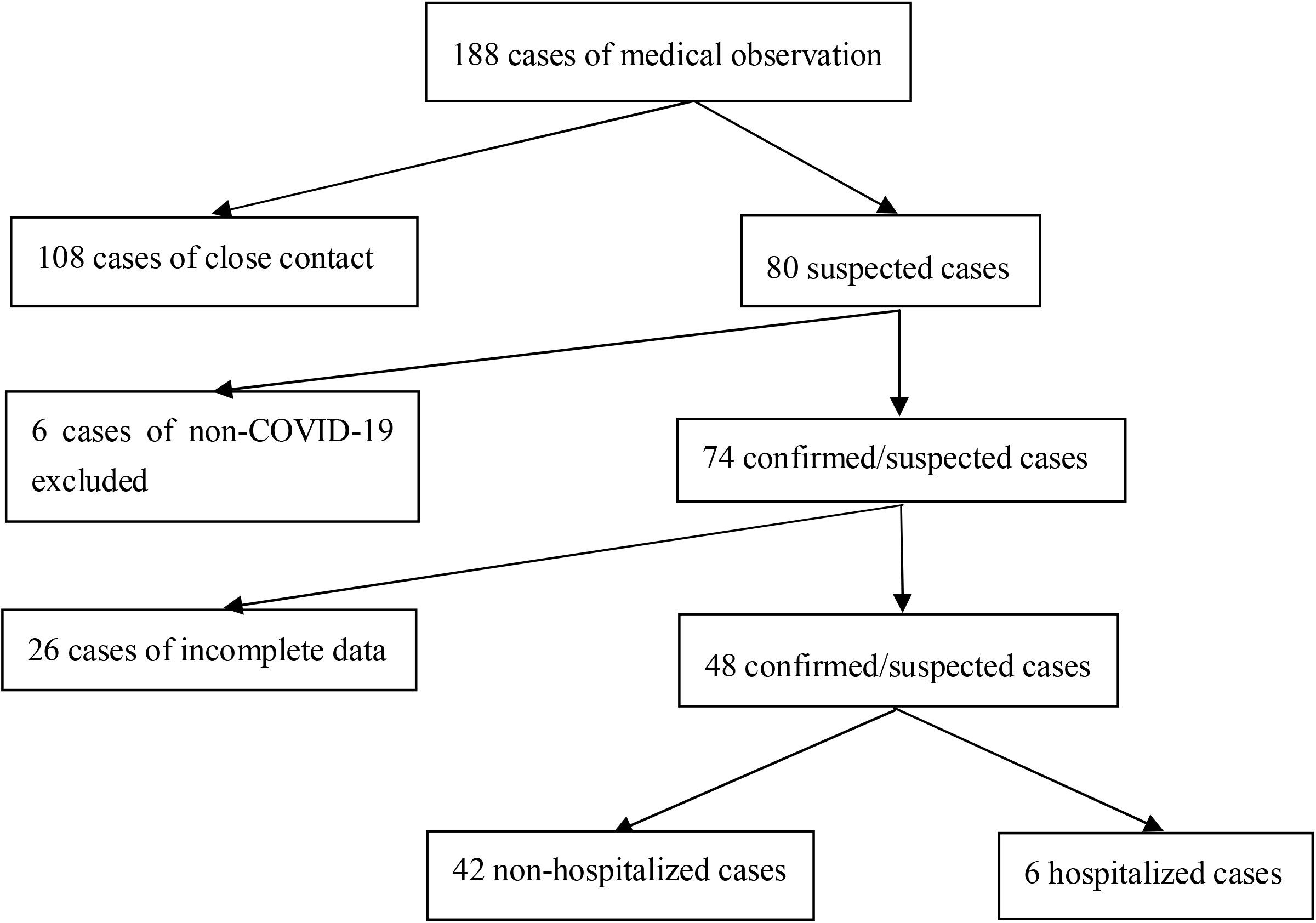
Patient enrollment flowchart.

**Figure 2:**
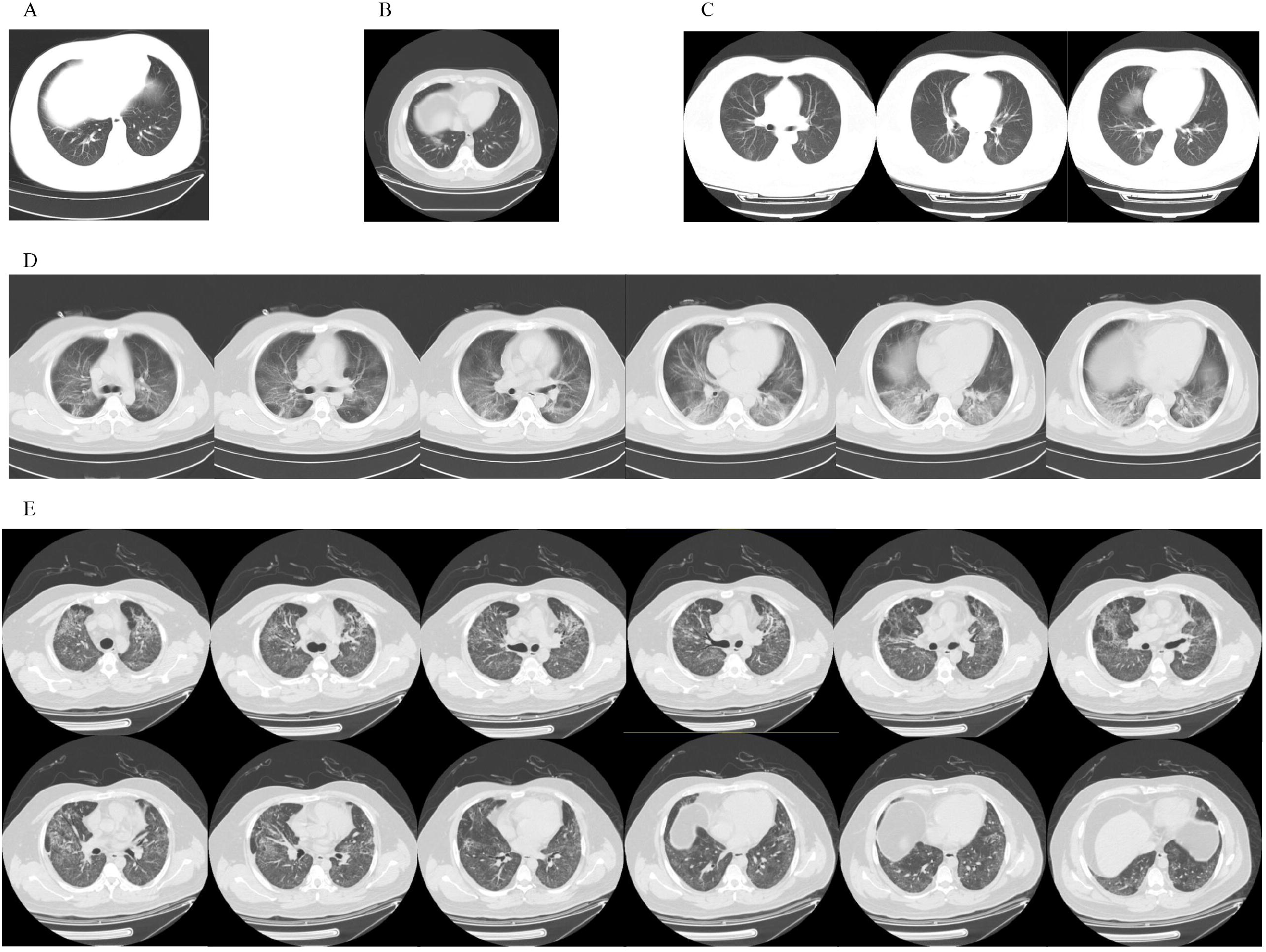
Chest CT images of patient 5. Transverse chest CT images showing patchy ground-glass opacities of the lower right lung on day 1 (A) and day 3 (B) after symptom onset. Transverse chest CT images showing bilateral ground-glass opacity on day 6 after symptom onset (C), and bilateral large infiltration shadow with partial consolidation on day 10 after symptom onset (D). On day 45 after symptom onset, transverse chest CT images showing bilateral lung diffuse reticular changes and fibrous stripes (E).

It is important to note that decreased lymphocyte count was observed in all inpatients, which is more prevalent than those previously reported. This difference may be attributed to the small sample size of our study.

### 3. Comparison of symptom trends between mild and severe cases

The hospitalized patients had significantly elevated body temperature, which scored between 3-4 points (around 38-40◻), that sustained longer than non-hospitalized patients. The mean body temperature of non-hospitalized patients with mild symptoms was below 38◻ on day 1, which continued to decrease and was back to normal by day 4-5 (figure 3A). We also found that hospitalized patients had elevated body temperature on day 5-6 along with exacerbated cough (figure 3B).

**Figure 3:**
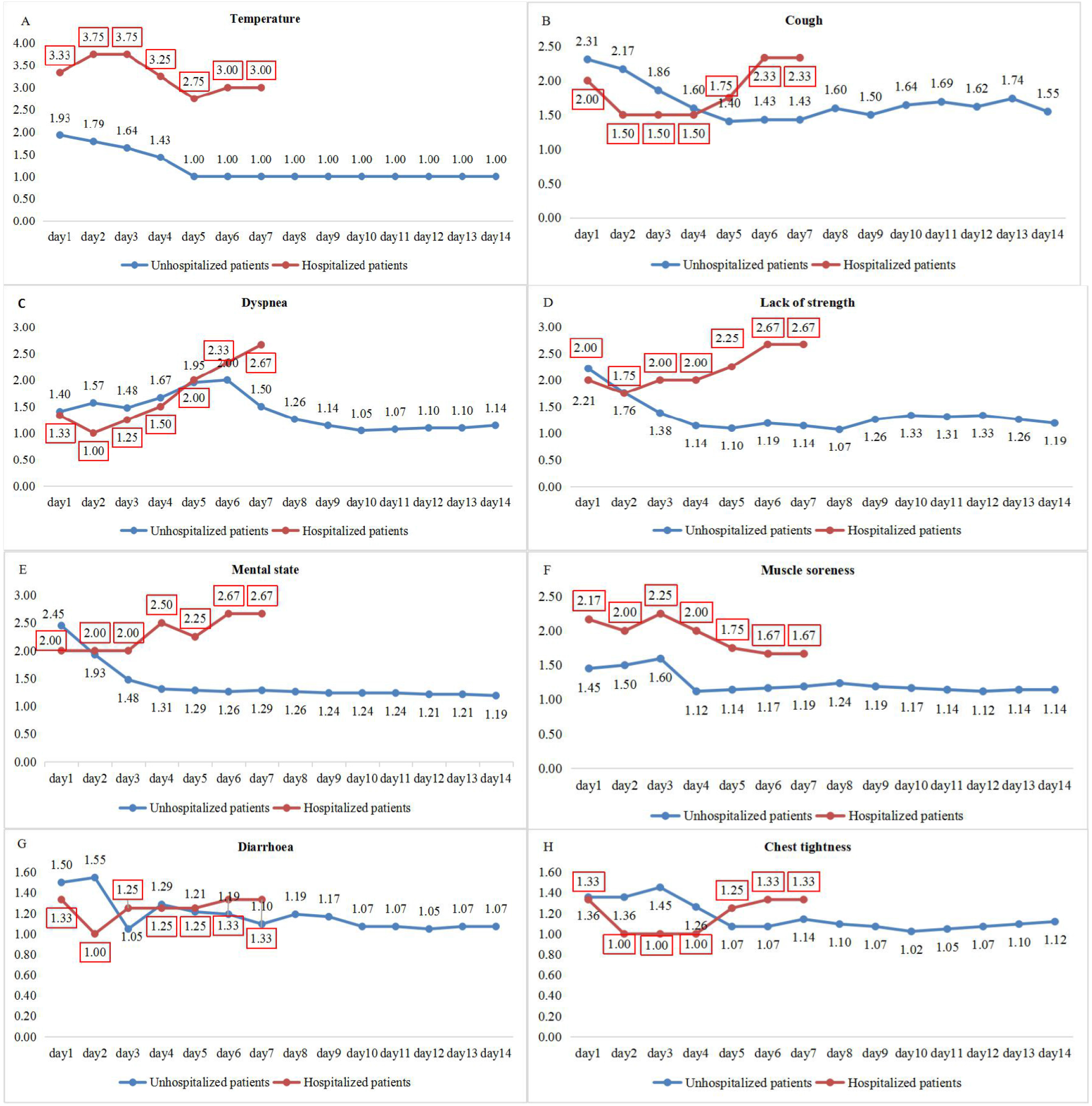
Comparison of symptom trends between unhospitalized and hospitalized patients.

The separation between the dyspnea curves was very distinct. For non-hospitalized patients with mild symptoms, dyspnea peaked on day 6 with a score of 2-3, which translates to shortness of breath when hurrying on a level surface. However, the symptom was gradually improved over time. In contrary, hospitalized patients had sustained dyspnea that continued to exacerbate over time(figure 3C).

We found that lack of strength, a symptom often neglected by clinical physicians, was a significant manifestation of COVID-19 patients. Although the degree of lack of strength seemed unrelated to disease severity at the onset of initial symptoms, this symptom was never alleviated in hospitalized patients and continued to exacerbate over time. On the other hand, this symptom was nearly gone on day 4 in non-hospitalized patients(figure 3D). In addition, we observed that the mental state of hospitalized patients gradually deteriorated over the course of disease, showing a similar progression as the lack of strength(figure 3E).

Furthermore, while muscle soreness was significantly alleviated on day 4 for both non-hospitalized and hospitalized patients(figure 3F), the progression of diarrhea was inconsistent for both groups of patients, which may be associated with the side effects of certain antiviral drugs(figure 3G).

## Discussion

The COVID-19 outbreak has hit all fever clinics of Wuhan’s class A tertiary hospitals harder than ever before and quarantine was the last resort we had under the impact of this PHEIC on our medical system. Given the current limited knowledge of the transmission pattern of COVID-19, the WHO emergency guidelines^9^ recommended that patients suspected of infection should be isolated and monitored in hospitals to ensure public health safety. Furthermore, in cases of insufficient hospitalization conditions, safety or medical resources, alternative quarantine methods, including home care and isolation, should be considered for suspected COVID-19 patients with mild symptoms. However, the guidelines did not provide clear instructions on how this should be conducted.

We used the O2O model and developed a new multidisciplinary home observation system led by medical staff from management, medicine, nursing, rehabilitation and psychology. This system uses WeChat and online document editing means to maintain continuous and interactive monitoring of the patients’ conditions in order to minimize the risk of cross-infection.

Among the 188 completed forms we collected, 108 were close contact cases without discomfort and clinical symptoms and were thereby removed from our study. In addition, 13·8% patients failed to complete the monitoring, and part of the reason is currently unknown. We suspect that it may be related to the setting of the form or the education level and lifestyle of the patients. Another reason for incompletion was that the patients only conducted less than 5 days of self-monitoring by the endpoint cut-off date of this study.

The gender ratio of our patients is different from those reported in recently published clinical data.^12-15^ The percentage of females was significantly higher in our study, which may be related to the fact that the scale was initially used by infected medical staff in our department, most of whom were nursing staff. This may also be the reason for the lower onset age we observed in our study.^12-16^ Nevertheless, we found that the hospitalized patients were significantly older than the non-hospitalized patients, which was consistent with previous findings.^12, 14^

We also had an interesting age-related observation in a father-daughter patient pair (patient 1 and patient 2). We found that although the father became ill first, the daughter had much severe symptoms except chest tightness and dyspnea than her father during the early stage of the disease. However, while the daughter’s symptoms gradually improved over time, the father’s chest tightness, dyspnea and previously latent symptoms worsened. Interestingly, the imaging features of these two patients were different from their respective clinical manifestations. The CT results of the daughter showed marked progression, whereas those of the father indicated minimal change. At the time of manuscript submission, the CT results of the daughter have significantly improved, and her symptoms have already disappeared. The patient is currently waiting for her second nucleic acid test results and may be discharged if the results turn out negative. On the other hand, the father is still experiencing mild dyspnea with no significant absorption by CT and must continue to be hospitalized for treatment. Based on the observations from this father-daughter patient pair, we speculate that elderly patients may have acute onset, early dyspnea, and symptoms that progress slow but are persistent and prone to turn critical. However, this speculation will need to be further confirmed by more samples.

All current studies have listed fever as an indispensable or highly prevalent symptom during the initial phase of COVID-19 infection.^12-15,17^ However, the study by Guan et al. showed that fever is only present in about half(48·7%) of the patients during initial diagnosis^16^, which is similar to our findings. Although fever was the most common initial symptom(58·3%) we observed in our patients, nasal congestion was also present in 6·3% of the patients, which was never reported in previous studies.^3-5,9-10,12-16^ One patient (2·1%) exhibited no symptoms during screening, and the CT results were negative for 10·4% of patients at initial diagnosis. Furthermore, the CT results were normal for 33·3% of hospitalized patients, which is consistent with the finding of Guan et al. that 20·9% of patients did not progress to pneumonia.^16^ Negative CT results not only make the clinical characteristics of patients seem more atypical, but they also increase the difficulty of initial screening.

We identified several indicators that were not significantly different between non-hospitalized and hospitalized patients (Table 1). Though, it is possible that the lack of differences in initial symptoms and laboratory test results between hospitalized and non-hospitalized patients is what makes it difficult for clinicians to identify patients with or prone to develop severe symptoms during the early stage.

Therefore, we believe that monitoring the trends of symptoms is more important for identifying severe cases and a unique finding of this study. Our study demonstrated that hospitalized patients had significantly elevated body temperature that sustained longer than non-hospitalized patients. In contrast, the body temperature of non-hospitalized patients with mild symptoms returned to normal by day 4-5. The separation in the dyspnea curves was also very distinct between the two groups of patients. While dyspnea peaked on day 6 for non-hospitalized patients with mild symptoms, it persisted in hospitalized patients and exacerbated over time. Similarly, the lack of strength and muscle soreness were both back to normal levels by day 4 for non-hospitalized patients. Whether these time points can serve as the turning points of the disease and whether medical staff should be alert to the possibility that the patients’ conditions may turn critical when the corresponding symptoms are not improved by these turning points will need to be further investigated.

The second unique finding of this study is that excessive laboratory data and physical examination are not necessary for the evaluation of patients with mild symptoms. Instead, the evaluation can be done through the patients’ subjective initiative and active participation in self-monitoring of the disease, which maximizes the reasonable allocation of medical resources. Therefore, our online evaluation system may prove useful in areas with insufficient medical resources.

In addition, the system we developed is the first to convert the subjective symptoms of patients into objective scores. This type of O2O, subjective-to-objective strategy may also be used in regions with similar highly infectious diseases to minimize the possibility of infection among medical staff. At the same time, the infection of family members was not increased during the strict self-isolation at home, so this proved that this isolated medical management mode was safe and effective. This is also consistent with the initial management model which was reported by Zhang et al in the Lancet Respir Med.^18^

This study has several limitations. First, our results were generated under a specific time and environment and were limited by the suddenness and complexity of the disease during the early stage, diversity and latency of the clinical manifestations, and the depletion of medical resources at the time. As a result, some data were incomplete and may result in bias in this study. Second, this study was a small sample survey of healthcare workers and their families. Aside from irreproducible interaction and discipline, the results obtained from this small sample may contain bias that need to be interpreted with caution. In addition, the 48 patients had no complications. Although the course of disease has shown a different trend between patients with mild and severe symptoms, it still had limitation. We will discuss it in further retrospective analysis of large samples in the future. Finally, we found later from our questionnaire that wearing a mask and keeping a distance of 1 meter, especially during home quarantine, were poorly accepted and inadequately executed by most patients(results to be published in another manuscript). Therefore, how to better implement these measures will need to be further pondered in subsequent work.

## Data Availability

After publication, the data will be made available to others on reasonable requests to the corresponding author. A proposal with detailed description of study objectives and statistical analysis plan will be needed for evaluation of the reasonability of requests. Additional materials might also be required during the process of evaluation. Deidentified participant data will be provided after approval from the corresponding author and Tongji Hospital.

## Contributors

SHuang, JD, YXiao and LA collected the epidemiological and clinical data. SLiu and BJ summarised all data. HX and LY drafted the manuscript. SLi revised the final manuscript.

## Declaration of interests

We declare no competing interests.

## Acknowledgments

We thank all patients and their families involved in the study.

## Notes

### Competing Interest Statement

The authors have declared no competing interest.

### Funding Statement

No funding.

## References

1 WHO. Statement on the second meeting of the International Health Regulations (2005) Emergency Committee regarding the outbreak of novel coronavirus (2019-nCoV). Jan 30, 2020. https://www.who.int/news-room/detail/30-01-2020-statement-on-the-second-meeting-of-the-international-health-regulations-(2005)-emergency-committee-regarding-the-outbreak-of-novel-coronavirus-(2019-ncov)

2 National Health Commission of the people’s Republic of China. Epidemic situation report. Feb 17, 2020. http://www.nhc.gov.cn/xcs/yqtb/202002/18546da875d74445bb537ab014e7a1c6.shtml

3 Holshue ML, DeBolt C, Lindquist S, et al. First Case of 2019 Novel Coronavirus in the United States. N Engl J Med 2020; published online Jan 31. DOI:10.1056/NEJMoa2001191.

4 Rothe C, Schunk M, Sothmann P, et al. Transmission of COVID-19 Infection from an Asymptomatic Contact in Germany. N Engl J Med 2020; published online Jan 30. DOI:10.1056/NEJMc200146.

5 Chan JFW, Yuan S, Kok KH, et al. A familial cluster of pneumonia associated with the 2019 novel coronavirus indicating person-to-person transmission: a study of a family cluster. Lancet 2020; published online Jan 24. https://doi.org/10.1016/S0140-6736(20)30154-9.

6 Li Q, Guan X, Wu P, et al. Early Transmission Dynamics in Wuhan, China, of Novel Coronavirus-Infected Pneumonia. N Engl J Med 2020; published online Jan 29. DOI: 10.1056/NEJMoa2001316.

7 Phan LT, Nguyen TV, Luong QC, et al. Importation and Human-to-Human Transmission of a Novel Coronavirus in Vietnam. N Engl J Med 2020; published online Jan 28. DOI:10.1056/NEJMc2001272

8 Cai JH, Zong WH. Analysis on the Application of Medical O2O Model in China. Chinese Journal of Health Information Management 2015. DOI: 10.3969/j.issn.1672-5166.2015.04.02

9 WHO. Home care for patients with suspected novel coronavirus (nCoV) infection presenting with mild symptoms and management of contacts. Feb 4, 2020. https://www.who.int/emergencies/diseases/novel-coronavirus-2019/technical-guidance/patient-management(accessed Feb 7, 2020)

10 National Health Commission of the people’s Republic of China. Diagnosis and treatment of pneumonia infected by the new novel coronavirus (the trial Fifth Edition). Feb 4, 2020. http://www.nhc.gov.cn/yzygj/s7653p/202002/3b09b894ac9b4204a79db5b8912d4440.shtml

11 Khamitov RA, Loginova S, Shchukina VN, Borisevich SV, Maksimov VA, Shuster AM. Antiviral activity of arbidol and its derivatives against the pathogen of severe acute respiratory syndrome in the cell cultures. Vopr Virusol 2008; 53: 9–13

12 Chen N, Zhou M, Dong X, et al. Epidemiological and clinical characteristics of 99 cases of 2019 novel coronavirus pneumonia in Wuhan, China: a descriptive study. Lancet 2020; 395(10223):507–513

13 Wang D, Hu B, Hu C, et al. Clinical Characteristics of 138 Hospitalized Patients With 2019 Novel Coronavirus-Infected Pneumonia in Wuhan, China. JAMA 2020; published online Feb 7. DOI: 10.1001/jama.2020.1585.

14 Huang C, Wang Y, Li X, et al. Clinical features of patients infected with 2019 novel coronavirus in Wuhan, China. Lancet 2020; published online Jan 24. https://doi.org/10.1016/S0140-6736(20)30183-5.

15 Chen L, Liu HG, Liu W, et al. Analysis of clinical characteristics of 29 cases of 2019-nCoV pneumonia. Chinese Journal of Tuberculosis and respiration 2020. DOI: 10.3760/cma.j.issn.1001-0939.2020.0005

16 Guan WJ, Ni ZY,Hu Y, et al. Clinical characteristics of 2019 novel coronavirus infection in China. medRxi 2020; published online Feb 9. http://dx.doi.org/10.1101/2020.02.06.20020974.

17 Zhu N,Zhang D, Wang W, et al. A Novel Coronavirus from Patients with Pneumonia in China, 2019. N Engl J Med 2020; published online Jan 24. DOI:10.1056/NEJMoa2001017.

18 Zhang J, Zhou L, et al. Therapeutic and triage strategies for 2019 novel coronavirus disease in fever clinics. Lancet Respir Med 2020; published online Feb 13. DOI: 10.1016/S2213-2600(20)30071-0.

